# Detection of persistent SARS-CoV-2 IgG antibodies in oral mucosal fluid and upper respiratory tract specimens following COVID-19 mRNA vaccination

**DOI:** 10.1101/2021.05.06.21256403

**Authors:** Aubree Mades, Prithivi Chellamathu, Lauren Lopez, Noah Kojima, Melanie A. MacMullan, Nicholas Denny, Aaron N. Angel, Joseph G. Casian, Matthew Brobeck, Nina Nirema, Jeffrey D Klausner, Frederick Turner, Vladimir I. Slepnev, Albina Ibrayeva

## Abstract

Previous studies have shown that mRNA COVID-19 vaccines are highly effective at preventing SAR-CoV-2 infection by generating an immune response, which in part produces SARS-CoV-2 IgG antibodies in serum. In this study, we hypothesized that COVID-19 vaccines may elicit production of SARS-CoV-2 IgG antibodies in the upper respiratory tract, such as in oral and nasal mucosal fluid. To test that hypothesis, we enrolled 114 participants within 3-7 days of receiving the first dose of the Moderna mRNA COVID-19 vaccine and collected oral mucosal fluid samples on days 5, 10, 15, and 20 after each vaccine dose. Of participants naive to SARS-CoV-2 (n = 89), 79 (85.4%) tested positive for SARS-CoV-2 IgG antibodies by time point 2 (10 days +/-2 days after first vaccine dose), and 100% tested positive for SARS-CoV-2 IgG by time point 3 (15 days +/-2 days after first vaccine dose). Additionally, we collected paired oral mucosal fluid and anterior nares samples from 10 participants who had received both vaccine doses. We found that participants had an average SARS-CoV-2 IgG antibody concentration of 2496.0 +/-2698.0ng/mL in nasal mucosal fluid versus 153.4 +/-141.0ng/mL in oral mucosal fluid. Here, we demonstrate detection and longitudinal persistence of SARS-CoV-2 IgG antibodies in upper respiratory tract specimens following COVID-19 mRNA vaccination.

## Introduction

Results from a phase 3 trial of the Moderna mRNA-1273 vaccine demonstrated 94% efficacy in preventing COVID-19 disease (1). Serum antibodies that were elicited by mRNA-1273 persisted through 6 months following the second vaccination dose, suggesting lasting protection against COVID-19 (2). We hypothesize that mRNA vaccines may elicit a strong antibody response in the upper respiratory tract at the sites where primary infection occurs and propagates, thereby preventing SARS-CoV-2 infection and transmission. An earlier work from our group demonstrated that SARS-CoV-2 IgG antibodies targeting spike protein S1 and S2 can be reliably detected from self-collected oral fluid among participants previously infected with SARS-CoV-2 (3). In the present study, we aimed to 1) assess whether SARS-CoV-2 IgG antibodies targeting the spike protein are detectable in self-collected oral and/or nasal mucosal following COVID-19 mRNA vaccination; and 2) quantify SARS-CoV-2 IgG antibodies in oral mucosal fluid over time in vaccinated participants and in unvaccinated participants following SARS-CoV-2 infection.

## Methods

Study enrollment was offered to healthcare workers at a COVID-19 testing site in San Dimas, California, USA. Enrollment was offered to subjects 18 years of age and older who had received the first dose of the COVID-19 Moderna vaccine within the previous 3-7 days, excluding subjects considered vulnerable, such as pregnant people, nursing home residents or other institutionalized persons, prisoners, and persons without decisional capacity. Verbal informed consent was obtained from each subject prior to enrollment in the study and any sample collection.

Enrolled participants provided oral fluid specimens on days 5, 10, 15, and 20 following their first vaccination dose, and days 5, 10, 15, and 20 following their second vaccination dose. At each time point, one sample was collected using the OraSure® Technologies Oral Specimen Collection Device (OSCD) for SARS-CoV-2 IgG antibody detection, and a second sample was collected using the Curative oral fluid swab test for SARS-CoV-2 RNA detection. Upon enrollment, participants were trained to self-collect specimens using each collection device while observed by study staff. Participants were provided additional test kits and asked to self-collect one of each specimen type unobserved at each follow-up time point, with a two-day grace period for sample collection and two day grace period for returning specimens to the study team.

63 days +/-5 days after the second COVID-19 vaccination dose, a sub-group of the cohort self-collected a paired oral and anterior mades specimen, using the OSCD and nasal swab, respectively, to assess the presence of SARS-CoV-2 IgG antibodies in the anterior nares mucosa following COVID-19 vaccination. Each collection device was weighed prior to and following specimen collection to measure the volume of sample collected. The dilution factor of each specimen was calculated by dividing the volume of preservative fluid (800 µL) by the volume of the sample. The final concentration of SARS-CoV-2 IgG antibodies was determined by multiplying the specimen’s determined antibody concentration by the calculated dilution factor. Antibody concentration for other specimens was not adjusted for the dilution factor.

Between April 2020 and December 2020, we collected oral fluid specimens using the OSCD from participants who were previously infected with SARS-CoV-2 in a separate study over time spanning a six month period. Here, we determined titers of SARS-CoV-2 IgG antibodies in these previously collected oral fluid specimens.

### OraSure® Technologies Oral Specimen Collection Device (OSCD) (item number 3001-2870, OraSure® Technologies, Bethlehem, PA)

The included cotton pad of the OSCD was brushed 5 times back and forth on each side of the lower and upper gums (lower left, lower right, upper left, and upper right gums for a total of 20 times) and then held stationary between the lower gum and the cheek on one side for 2 to 5 min. The pad was then placed into the provided tub, containing 800µL of preservative fluid. The cotton pad collects approximately 800 µL of oral mucosal fluid. Specimens were left at room temperature for up to 5 days and then stored at -80°C until use.

### Anterior Nares Swab (item number CY-98000; Huachenyang Technology, China)

A flocked nylon swab was swirled around each anterior nare for ten seconds and then placed into the tube provided with the OSCD. The swab collects approximately 25 µL of nasal fluid and the provided tube contains 800 µL of preservative fluid. Specimens were left at room temperature for up to 5 days and then stored at -80°C until use.

### Quantitative SARS-CoV-2 IgG ELISA Assay

A quantitative SARS-CoV-2 IgG ELISA was performed on self-collected oral mucosa specimens collected using the OSCD and anterior nares specimens using the flocked swab. All reagents and proteins were obtained from OraSure® Technologies. 25 μL of sample diluent and 100 µL of either oral or nasal mucosa specimen were added to 96-well plates coated with both S1 and S2 subunits of the SARS-CoV-2 viral spike glycoprotein. Plates were incubated at ambient temperature for 1h. Sample wells were then washed six times with wash buffer (20x dilution with ddH2O, 350 µL per well) and conjugate solution was added (100 µL□per well). Plates were incubated at ambient temperature for 1□h and sample wells were then washed an additional six times. Next, substrate solution was added (100□µL per well) and the plate was incubated at ambient temperature for 30 min. Finally, stop solution was added (100 µL per well). The absorbance of sample wells was measured immediately at 450□nm and 630□nm. Output reports generated the absorbance at 630□nm subtracted from the absorbance at 450□nm.

To quantify SARS-CoV-2 IgG antibodies in oral mucosal fluid and nasal samples, an S1-specific monoclonal IgG antibody with no known cross-reactivity to the S2 domain of the spike protein was used as a reference antibody. Cross-reactivity from IgA and IgM were minimal as reported by the manufacturer but was also tested. We confirmed the lack of cross-reactivity from IgA with the absence of detectable IgA signal on an assay. A standard curve was developed using a monoclonal IgG antibody targeting the S1 antigen of SARS-CoV-2 at concentrations of 0, 1.5, 3, 6, 12, and 20ng/mL with a polynomial regression curve-fitting model. The standard curve was used to calculate the sample IgG antibody concentration from absorbance values at 450/630 nm from the ELISA assay. The absorbance signal from each sample is directly proportional to the IgG antibody concentration present in the oral fluid (**Fig. 1**). Specimens with antibody titer levels exceeding the range of the standard curve were diluted in a sample dilution buffer and re-ran.

**Figure 1.**
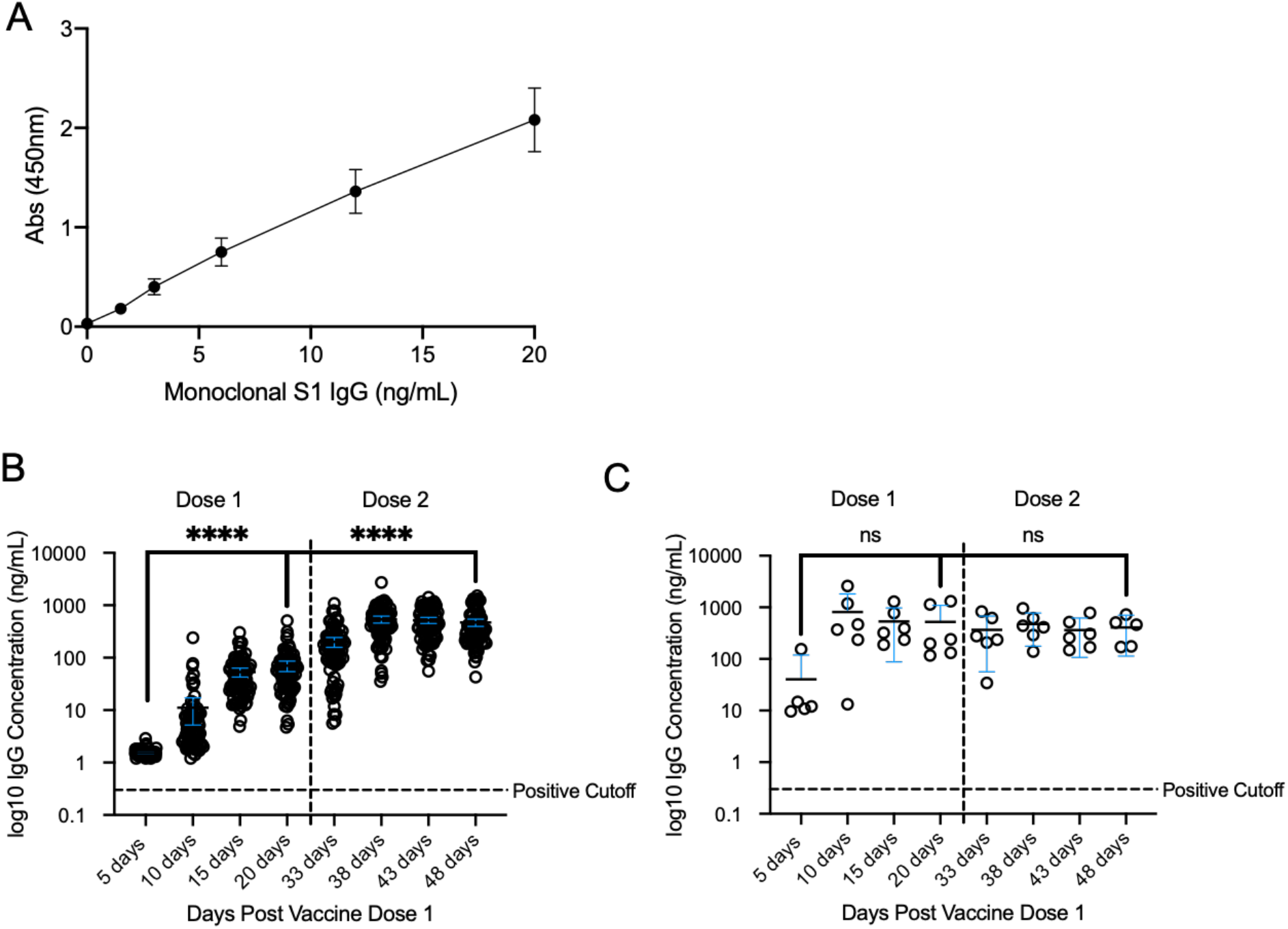
Quantification of IgG antibodies against SARS-CoV-2 in oral mucosa in participants with (n = 6) and without (n = 89) prior SARS-CoV-2 infection that were vaccinated by the Moderna COVID-19 vaccine. **A)** Monoclonal IgG targeting the S1 antigen of SARS-CoV-2 was used to develop a quantitative assay for antibody quantification. The standard curve was developed based on monoclonal IgG antibody concentrations of 0ng/mL, 1.5ng/mL, 3ng/mL, 6ng/mL, 12ng/mL and 20ng/mL. The expression obtained from this standard curve, y = -0.00121×2 + 0.128x + 0.0163, was used to determine sample concentrations from optical density (O.D.) measured at 450nm. **B)** Antibody quantitation in COVID-naive individuals (n = 89) with a 95% confidence interval (CI). 20 days after the first vaccine dose, the average antibody concentration among individuals was 70.25 +/-73.0ng/mL while 20 days after the second vaccine dose, the average antibody concentration among individuals was 470.4 +/-352.2ng/mL. The difference between means across collected timepoints was found to be significant (p < 0.0001) between 5 and 20 days and 20 and 48 days. **C)** Antibody quantification in individuals previously infected by SARS-CoV-2 (n = 6) with a 95% CI. 20 days after the first vaccine dose, the average antibody concentration among individuals was 523.1+/-548.6ng/mL while 20 days after the second vaccine dose, the average antibody concentration among individuals was 409.1 +/-237.4ng/mL.

The Limit of Detection (LoD) was 1 ng/mL and the Limit of Quantification (LoQ) was 1.5 ng/mL. The calculated values for oral fluid specimens presented in this paper represent the diluted specimen, with approximately 800 µL of oral fluid diluted in 800 µL preservative. The real oral fluid IgG antibody concentration is up to several folds larger than the value reported, depending on the volume of fluid collected.

### Curative Real-Time rRT-PCR SARS-CoV-2 Collection and Assay

Oral fluid specimens were collected according to the Curative Instructions for Use as described in the FDA-EUA rRT-PCR SARS-CoV-2 Assay which includes expectoration of upper respiratory tract fluid (i.e., 3-5 coughs). Specimens were analyzed for SARS-CoV-2 RNA using the Curative FDA-EUA oral fluid COVID-19 test.

### Data Analysis

All data analysis was performed using GraphPad Prism (GraphPad Prism Version 9.0.2) software. Paired, two-sided t-tests were performed to compare differences in average antibody concentrations across conditions. All data can be made available upon request.

## Ethics

The study protocols were approved by the Advarra Institutional Review Board (IRB# Pro00048737 and Pro00045766).

## Results

In January 2021, we enrolled 114 participants who had received the Moderna mRNA COVID vaccine in the previous 3-7 days. During the study period, 18 participants were excluded from the study due to incomplete sample return. Of the remaining 96 participants, the average participant age was 29.6 years (interquartile range [IQR: 24.3, 30.5 years]). 61 (63.5%) identified as female, and 6 (6.3%) had tested positive for SARS-CoV-2 RNA at some time prior to the study. 1 participant had a previous SARS-CoV-2 infection detected and confirmed by rRT-PCR, but tested negative for SARS-CoV-2 IgG antibodies in both serum and oral specimens prior to COVID-19 mRNA vaccination. This participant was not included in the cohort of participants with previous COVID-19 infection, but developed antibodies by time point 2 (10 days +/-2 days following first vaccination dose) and had a SARS-CoV-2 IgG antibody concentration of 125.57ng/mL at time point 8 (20 days +/-2 days following second vaccination dose). To the best of our knowledge, the remaining 89 participants were naive to SARS-CoV-2 prior to the study.

Ninety-five participants (99.0%) tested negative for SARS-CoV-2 RNA at every time point throughout the study. 1 participant (1.0%) tested positive for SARS-CoV-2 RNA (on days 5 and 10 after receiving the second dose of the Moderna vaccine, with a SARS-CoV-2 CT value of 28.77 and 31.42, respectively). This participant did not develop severe symptoms to COVID-19.

Reviewing vaccination records and testing results of all healthcare workers at the COVID-19 testing site where enrollment occurred throughout the study period, we observed 0.60% prevalence of SARS-CoV-2 infection in employees who had received both doses of a COVID-19 vaccine, and 5.94% prevalence in unvaccinated employees.

Of participants naive to SARS-CoV-2 (n = 89), 79 (88.8%) tested positive for oral mucosa SARS-CoV-2 IgG antibodies by time point 2 (10 days +/-2 days after first vaccine dose), and 100% tested positive for oral fluid SARS-CoV-2 IgG by time point 3 (15 days +/-2 days after first vaccine dose).

For this group, the average oral fluid IgG antibody concentration 20 days after the first vaccine dose (70.25 +/-72.95ng/mL) was found to be significantly higher (p < 0.0001) than the average oral concentration at 5 days (1.517 +/-0.25ng/mL). Further, the average concentration 20 days after the second vaccine dose (470.4 +/-352.2ng/mL) was found to be significantly higher (p < 0.0001) than the antibody concentration observed 20 days after the first vaccine dose.

Of the 6 participants who were previously infected with SARS-CoV-2, 6 (100%) tested positive for SARS-CoV-2 IgG antibodies at time point 1 (5 days +/-2 days). For this group, the average concentration 20 days after the first vaccine dose was 523.1+/-548.6ng/mL and the average concentration 20 days after the second dose was 409.1 +/-237.4ng/mL.

### Paired oral and nasal samples

In the subgroup of participants that provided a paired sample of anterior nares swab specimen and OSCD oral fluid specimen following their second vaccination dose (n=10), 100% of the anterior nare mucosa specimens had detectable antibodies with an average IgG antibody concentration of 2496.0 +/-2698.0 ng/mL when adjusted for individual dilution factor. Paired oral mucosa samples had an average IgG antibody concentration of 153.4 +/-141.0 ng/mL when adjusted for individual dilution factor. A paired two-tailed t-test revealed a significant difference (p-value = 0.0232) in mean IgG values between SARS-CoV-2 IgG antibody concentrations in each specimen type (**Fig. 2**)

**Figure 2.**
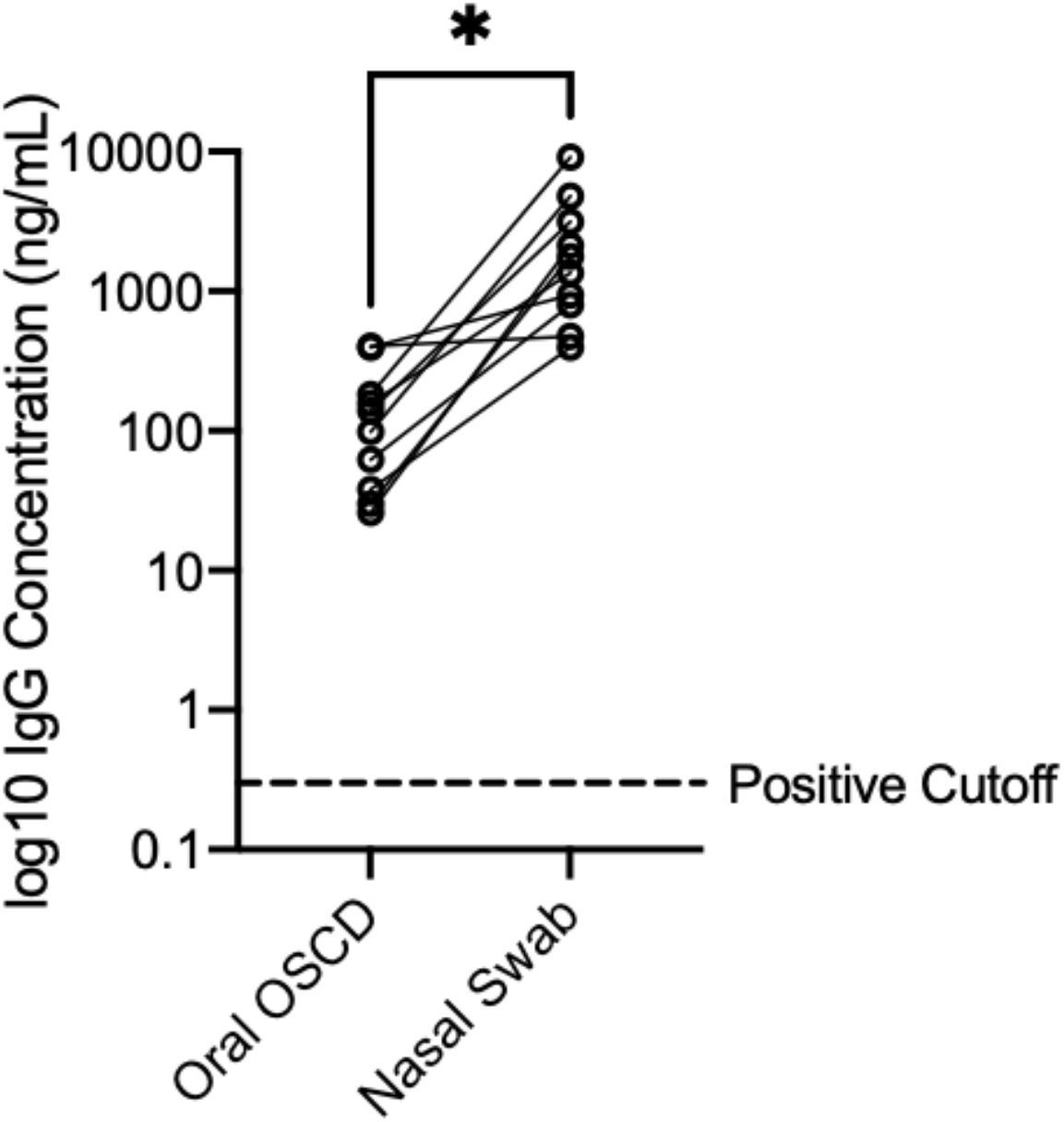
Antibody quantification in paired specimens collected by both anterior nares swabs and oral OSCDs reveal a higher antibody concentration in the nasal passage. After receiving both doses of the Moderna vaccine, all individuals (n = 10) had detectable antibodies collected from their anterior nares with an average concentration of 2496.0 +/-2698.0ng/mL in nasal fluid. The same individuals had a mean concentration of 153.4 +/-141.0ng/mL in saliva collected using the Oral OSCD. Concentration values were adjusted for sample collection and diluent volumes. The difference between mean antibody concentrations from different collection methods was found to be significant using a paired two-tailed t-test (p = 0.0232).

### Participants with prior SARS-CoV-2 infection not vaccinated against COVID-19

Self-collected oral fluid samples using the OSCD from the cohort of unvaccinated participants with a previous SARS-CoV-2 infection (n = 31) were evaluated over time (**Fig. 3**) 116 +/-14 days following infection, the average concentration of SARS-CoV-2 IgG antibodies among these individuals was found to be 23.7 +/-22.5 ng/mL.

**Figure 3.**
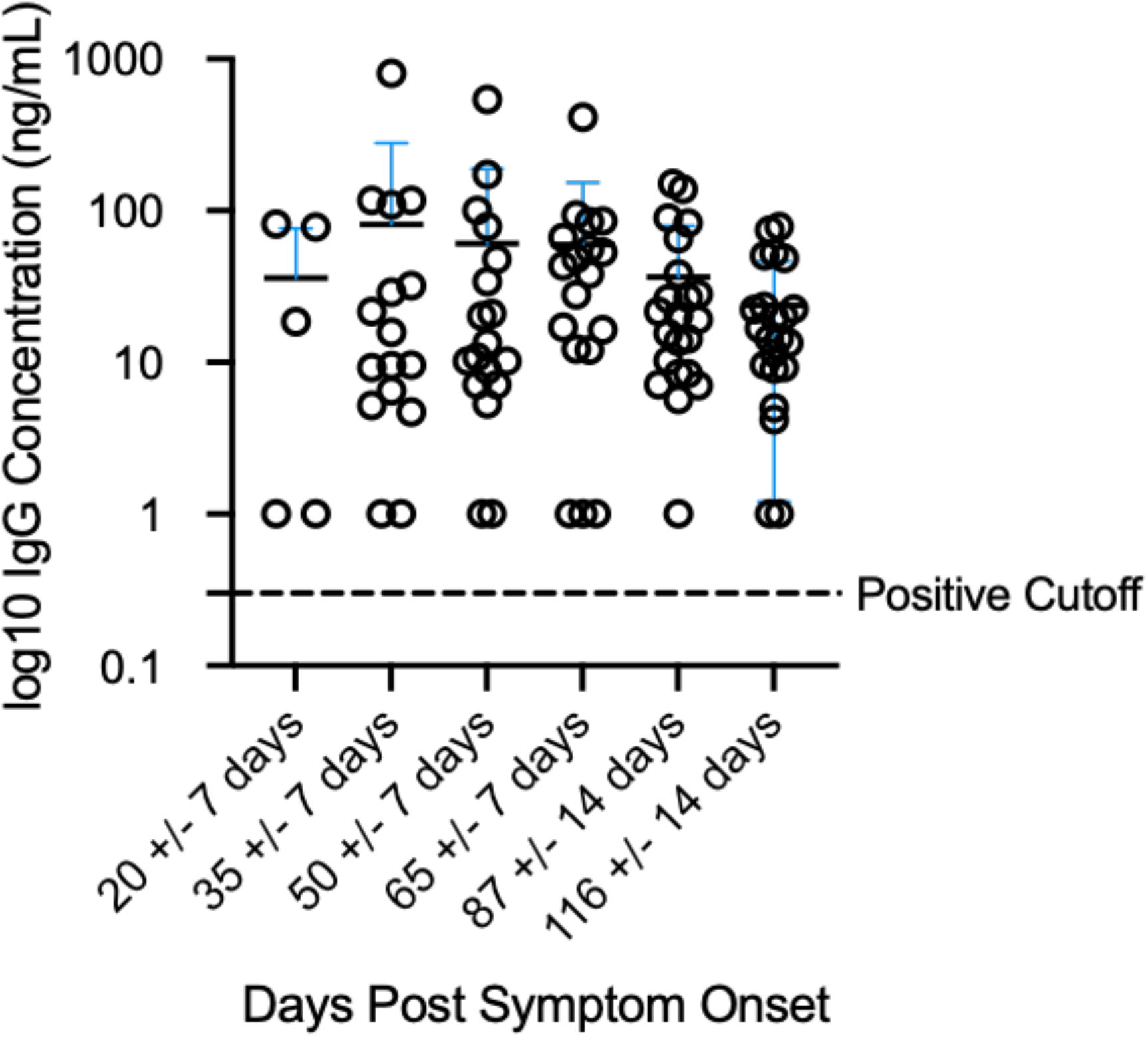
Quantification of IgG antibodies against SARS-CoV-2 in oral fluid samples collected from individuals (n = 31) infected by SARS-CoV-2 in the months following symptom onset. 116 +/-14 days post symptom onset, individuals had an average SARS-CoV-2 S1 protein concentration of 23.7 +/-22.5 ng/mL.

## Discussion

To our knowledge, this is the first demonstration that SARS-CoV-2 IgG antibodies can be detected and quantified in self-collected oral fluid and nasal swab specimens following vaccination with a COVID mRNA vaccine. We detected SARS-CoV-2 IgG antibodies in 100% of self-collected oral fluid specimens from participants by 15 days +/-2 days after the first vaccination dose. Our observations are in agreement with the Moderna vaccine clinical study, which determined that among adults aged 18 to 70 years of age, SARS-CoV-2 IgG antibodies were detected among all participants who received a COVID-19 vaccination series in serum samples by day 15 (4–7).

In vaccinated participants, the average antibody concentration increased significantly from 70.25 ng/mL on time point 4 (20 days +/-2 days after the first dose), to 470.4 ng/mL on time point 8 (20 +/-2 days days after the second vaccine dose). Taking account of variation in sample dilution, we expect the real antibody concentration to be 1.5-2-fold higher than these calculated values. Significantly higher SARS-CoV-2 IgG antibody concentrations were observed in anterior nares specimens compared to OSCD oral fluid specimens, after adjusting for dilution factor.

Of vaccinated participants with a prior infection of SARS-CoV-2 (n=6), all participants (100%) had detectable SARS-CoV-2 IgG antibodies by time point 1 (5 days +/-2 days after the first vaccination dose). Because we did not collect participant samples prior to time point 1, it is not known how many of those participants had SARS-CoV-2 IgG antibodies prior to vaccination. In this cohort, there were too few individuals to determine whether there was a significant difference in average SARS-CoV-2 IgG antibody titers at any time point following both vaccination doses.

This study had several limitations. Due to our enrollment of health care workers, the average age of participants was low, with only 8 participants over the age of 50 years and only 1 participant over the age of 65 years. Participants were not monitored during specimen collection after the first time point, but the ability of participants to follow instructions for unobserved collection was validated internally prior to sample collection. All participants received the Moderna COVID-19 vaccine, and further research is needed to understand the longitudinal antibody response in oral or nasal mucosal fluid following other COVID-19 vaccines, such as the mRNA Pfizer/BioNtech vaccine and the viral vector Janssen vaccine. It is not yet known how oral mucosal fluid antibody titers are directly comparable to serum titers for the protection of infection or disease. Self-collected oral mucosal fluid specimens for SARS-CoV-2 quantification could have potential applications in monitoring population antibody seroprevalence and long-term immune response to COVID-19 vaccines.

Quantification of SARS-CoV-2 IgG antibodies in oral fluid mucosal specimens of non-vaccinated individuals with previous SARS-CoV-2 infection demonstrated antibody persistence through 3.5 months without a significant decline. This may suggest persistence of SARS-CoV-2 IgG antibodies in oral mucosal fluid post-vaccination.

## Conclusions

To the best of our knowledge, this is the first demonstration of persistent IgG antibody presence in oral and nasal mucosa following COVID-19 vaccination with a COVID-19 mRNA vaccine.

High antibody concentration at the sites of the primary infection may play a direct role in preventing viral transmission. Additional *in-vitro* experiments to study the effects of treating SARS-CoV-2 viral cultures with oral mucosal samples from vaccinated individuals would be required to test this hypothesis.

All 95 participants (100%) who received a COVID-19 mRNA vaccination had detectable SARS-CoV-2 IgG antibodies in oral mucosa fluid by 15 days +/-2 days after their first COVID-19 vaccine dose. On average, participants with no prior history of SARS-CoV-2 infection developed a higher average antibody concentration after the second vaccine dose. Participants with a history of COVID-19 experienced a greater initial antibody titer, and greater titers at each time point, in comparison to participants naive to SARS-CoV-2, but no significant change in antibody titer following their second vaccination dose. Further research is needed to characterize how SARS-CoV-2 respiratory IgG antibody titers are elicited by different commercially available

COVID-19 vaccines, among older adults, and over a longer study period, to monitor the scale and the duration of the protection against COVID-19 infection via IgG antibodies.

Testing of the respiratory specimens for SARS-CoV-2 IgG antibody offers a self-collected, non-invasive specimen collection alternative to serum specimens that can be scaled with a higher expected patient compliance.

## Data Availability

Data is available upon request from Corresponding Author

## Author Contributions

A.M. designed the study protocol, designed experiments, analyzed and interpreted data, and drafted the manuscript. P.C. designed and ran experiments, analyzed and interpreted data, and drafted the manuscript. L.L., M.B., and N.N. designed the study protocol and sample collection and edited the manuscript. N.K. and J.D.K. provided medical oversight, conceptualized and designed the study protocol, and edited the manuscript. M.A.M analyzed and interpreted data and edited the manuscript. N.D., A.N.A., and J.G.C. ran experiments. F.T. and V.I.S conceptualized and designed the study protocol, designed experiments, and edited the manuscript. A.I. oversaw and designed experiments, oversaw data collection and analysis, and drafted the manuscript.

## Declarations

All authors are, or were at the time of research, employed by Curative Inc., a diagnostics company. F.T. and V.S. are shareholders of Curative Inc. Curative Inc. does not have conflicts of interest with OraSure® Technologies to disclose.

## Acknowledgements

We thank Ronell Lopez, Marilisa Santacruz, Joseph Kapcia, Colin Witt, Cedie Bagos, Christina Le, Alexis Alexander, and Stephanie Zhang from Curative Inc.

## Keywords

OSCD: OraSure® Technologies Oral Specimen Collection Device
rRT-PCR: Real-time reverse transcription polymerase chain reaction
ELISA: Enzyme-Linked Immunosorbent Assay
COVID-19: 2019 novel coronavirus
RNA: ribonucleic acid
mRNA: Messenger ribonucleic acid
IgA: Immunoglobulin A
IgG: Immunoglobulin G
IgM: Immunoglobulin M
SARS-CoV-2: Severe acute respiratory syndrome coronavirus 2
LOD: Limit of Detection
LOQ: Limit of Quantification

